# Effects of CFTR Modulators on Serum Biomarkers of Liver Fibrosis in Children with Cystic Fibrosis

**DOI:** 10.1101/2022.09.02.22279547

**Authors:** Steven Levitte, Yonathan Fuchs, Russell Wise, Zachary M. Sellers

**Affiliations:** Division of Pediatric Gastroenterology, Hepatology, and Nutrition, Stanford University, Palo Alto, CA 94304, USA; Department of Pharmacy, Lucile Packard Children’s Hospital Stanford, Palo Alto, CA, 94304, USA

**Keywords:** CF, CFTR, modulators, liver function tests, liver disease

## Abstract

The cystic fibrosis transmembrane conductance regulator (CFTR) corrector/potentiator combinations lumacaftor/ivacaftor and elexacaftor/tezacaftor/ivacaftor improve sweat chloride, pulmonary function, and nutrition. Yet it is unclear whether they may also impact the progression of liver fibrosis, which is a substantial source of morbidity and mortality for patients with cystic fibrosis (CF). We conducted a retrospective, single-center analysis of children and adolescents with CF treated with lumacaftor/ivacaftor and/or elexacaftor/tezacaftor/ivacaftor therapy, focusing on alterations in liver function tests and fibrosis indices using previously-established thresholds that corresponded with increased liver elastography. In pairwise comparisons of before and during treatment timepoints, we found that those with CF-associated liver involvement experienced significant decreases in gamma-glutamyl transferase (GGT), AST-to-Platelet Index (APRI), and GGT-to-Platelet Ratio (GPR) while on lumacaftor/ivacaftor. These differences were not observed in patients treated with elexacaftor/tezacaftor/ivacaftor, nor were they observed in patients without underlying CF-associated liver disease. These results provide the first evidence that lumacaftor/ivacaftor may improve liver fibrosis in children and adolescents with CF and suggest it may be beneficial in the treatment of CF-associated liver disease.

## INTRODUCTION

Lumacaftor/ivacaftor and elexacaftor/tezacaftor/ivacaftor are approved in the United States for the treatment of cystic fibrosis (CF) in those with homozygous or compound heterozygous F508del cystic fibrosis transmembrane conductance regulator (CFTR) mutations, respectively. Clinical trials showed that these CFTR corrector/potentiator combination drugs improve pulmonary outcomes (1, 2); however, there is little data on the impact of these therapies on liver health. This reflects a major unmet clinical need, as liver complications remain a leading cause of death in patients with CF (3). We and others have previously shown that gamma-glutamyl transferase (GGT), platelet levels, and the liver fibrosis indices APRI (AST-to-Platelet Ratio Index) and GPR (GGT-to-Platelet Ratio) correlate with imaging-based markers of liver fibrosis and cirrhosis in CF (4-6). We used these surrogates for liver fibrosis to evaluate the impact of lumacaftor/ivacaftor and elexacaftor/tezacaftor/ivacaftor on children and adolescents with CF with and without abnormal liver fibrosis indices.

## METHODS

### Human Subjects

Retrospective analysis of all patients cared for at a tertiary care CF Foundation-accredited academic Pediatric CF Center between January 1, 2013 and December 1, 2021 was undertaken with Institutional Review Board approval (#11197). Inclusion criteria were: 1) lumacaftor/ivacaftor or elexacaftor/tezacaftor/ivacaftor for ≥ 6 months, 2) ≥ 1 set of liver function tests and platelets prior to and during treatment. We included data from six patients who participated in open-label extension studies of NCT02514473 and/or NCT03061331; one patient participated in the randomized control trial NCT02514473 with unknown randomization, and was excluded (7, 8). Exclusion criteria included liver transplantation. We separated patients based on “liver involvement” or “no liver involvement” prior to starting lumacaftor/ivacaftor or elexacaftor/tezacaftor/ivacaftor. Liver involvement was defined as APRI ? 0.345 or GPR ? 0.403 on at least two occasions at least three months apart prior to starting CFTR modulator therapy. These cut-offs were previously validated to detect liver fibrosis in CF using ultrasound and magnetic resonance elastography (4, 9).

### Liver function tests and liver fibrosis indices

All available laboratory tests were examined during the aforementioned period with focused attention on aspartate aminotransferase (AST), alanine aminotransferase (ALT), GGT, and platelets. For pre-treatment, those labs closest to and before the start of lumacaftor/ivacaftor were used. For post-treatment the most recent labs were used for analysis, with the exception of seven patients who were treated with lumacaftor/ivacaftor followed by elexacaftor/tezacaftor/ivacaftor. For these patients, laboratory values while on lumacaftor/ivacaftor were examined as close as possible to the treatment time on elexacaftor/tezacaftor/ivacaftor. Upper limits of normal (ULN) for AST, ALT, and GGT values were based on Canadian Laboratory Initiative on Pediatric Reference Intervals (CALIPER), which accounts for sex and age (10). Platelet values of 150 x10^3^/µL were used as the lower limit of normal (LLN). Laboratory values were excluded if associated with inpatient admissions, documented infections, or were deemed secondary to other drug treatments based on clinical documentation review by a physician (SL) and a pharmacist (RW). All laboratory values >2 times the ULN were evaluated to determine if they occurred within 3 months of the patient taking any of the following medications: ciprofloxacin, clarithromycin, erythromycin, ethambutol, fluconazole, itraconazole, levofloxacin, minocycline, posaconazole, and voriconazole. More extensive temporal analysis was performed with values >3 times the ULN, which identified drug-related elevations secondary to amoxicillin/clavulanic acid, sulfamethoxazole/trimethoprim, and tobramycin. APRI and GPR were calculated using CALIPER ULN values using the following equation:

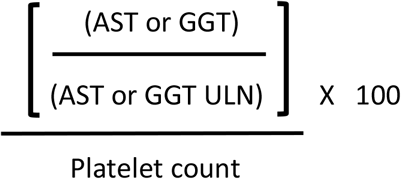

### Statistics

Mean, median, and standard deviation (SD) were calculated. Graphs were generated using Prism9 (GraphPad Software, San Diego, CA). Significance was determined by P values <0.05 using two-tailed paired nonparametric Wilcoxon t-test for pre- and post-treatment analysis. Chi-squared test was used to evaluate differences in proportions of patients where indicated.

## RESULTS

*Cohorts*. Of the approximately 230 children and adolescents at our Pediatric CF Center we identified 42 patients that were treated with lumacaftor/ivacaftor and 52 that were treated with elexacaftor/tezacaftor/ivacaftor for ≥6 months. Ten patients were excluded due to unavailable laboratory values, resulting in 84 patients (35 lumacaftor/ivacaftor, 49 elexacaftor/tezacaftor/ivacaftor) for analysis. Thirteen of 35 in the lumacaftor/ivacaftor group and 15/49 in the elexacaftor/tezacaftor/ivacaftor group had liver involvement prior to starting CFTR modulator therapy (Table 1). Among these, seven patients were treated with lumacaftor/ivacaftor followed by elexacaftor/lumacaftor/ivacaftor and had liver involvement before starting lumacaftor/ivacaftor (Table 1).

**Table 1.**
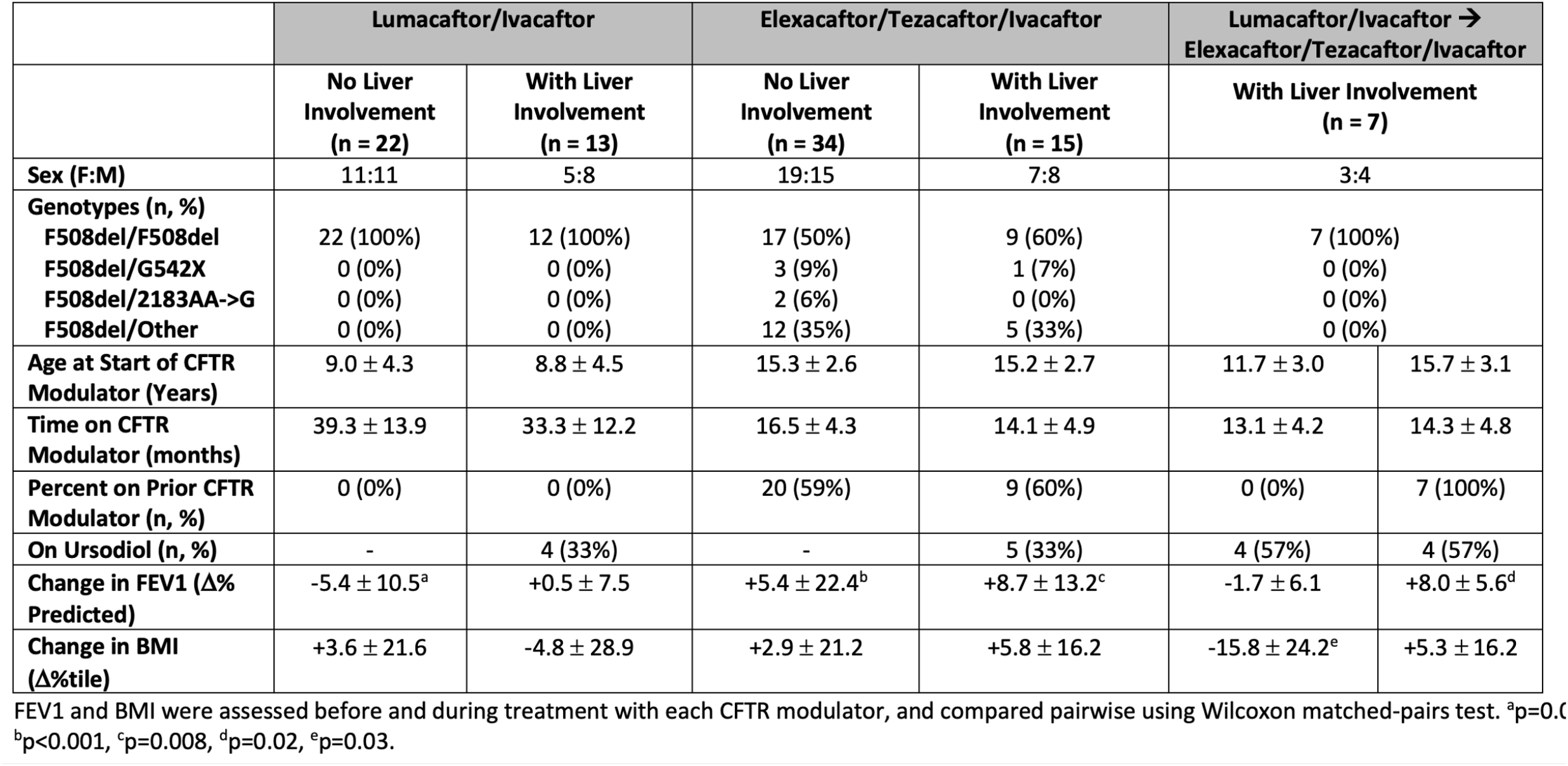
Characteristics of Cohorts.

### Liver function tests and fibrosis indices in those with and without pre-existing liver abnormalities

In order to determine if lumacaftor/ivacaftor and/or elexacaftor/tezacaftor/ivacaftor improves liver outcomes in CF, we examined LFTs and liver fibrosis indices separately in those patients with and without CF-associated liver involvement. We compared values obtained from the same patients prior to starting lumacaftor/ivacaftor with laboratory values obtained after an average (SD) of 33.3 ± 12.2 months of therapy for patients with liver involvement and 39.3 ± 13.9 months in patients without liver involvement. In patients with underlying liver involvement, we observed a significant decrease in GGT, APRI, and GPR (P = 0.002, P = 0.042 P = 0.002, respectively, Figure 1C, 1E, and 1F). We did not observe significant differences in AST, ALT, or Platelets (Figure 1A, 1B, and 1D). Separately, we performed these same analyses on patients who did not have documented liver involvement prior to starting lumacaftor/ivacaftor. We did not observe a significant difference in any of the LFTs or liver fibrosis indices (Supplemental Figure 1A-F). We performed the same analysis in patients treated with elexacaftor/tezacaftor/ivacaftor (average (SD) of 14.1 ± 4.9 months for those with liver involvement and 16.5 ± 4.3 months for those without liver involvement). In those with liver involvement, we observed an increase in ALT (P = 0.048, Figure 1H) and a decrease in platelets (P = 0.010, Figure 1J). We did not observe significant alterations in AST, GGT, APRI, or GPR (Figure 1G, 1I, 1K, and 1L). In patients without liver involvement treated with elexacaftor/tezacaftor/ivacaftor, we observed increases in ALT, APRI, and GPR (P = 0.005, P = 0.027, and P = 0.023, respectively, Supplemental Figure 1H, 1K, and 1L) and a decrease in platelets (P = 0.002, Supplemental Figure 1J). GGT trended towards an increase although this was not significant (P = 0.063, Supplemental Figure 1I).

**Figure 1.**
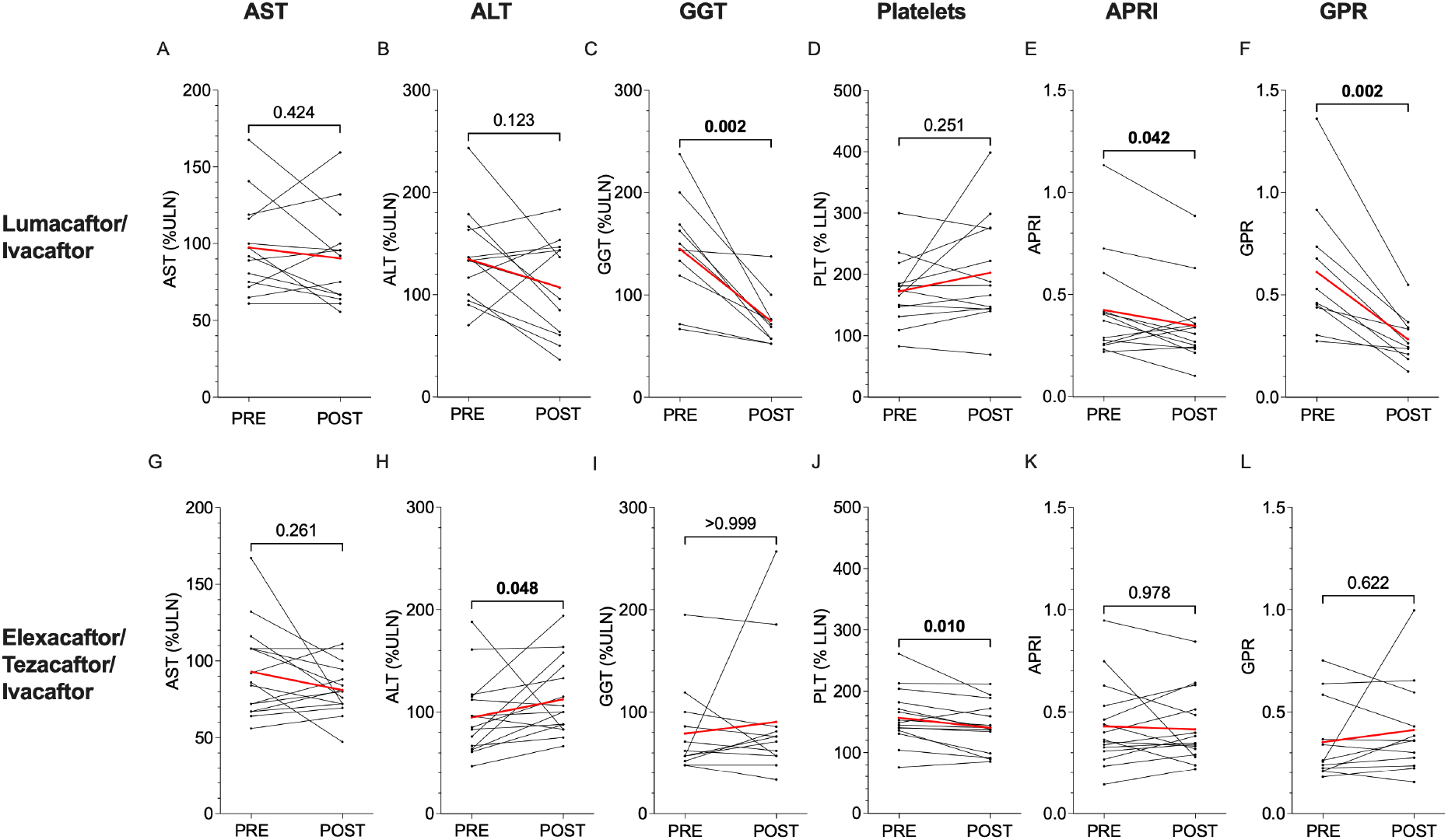
Changes in Liver Function Tests and Liver Fibrosis Indices During Lumacaftor/Ivacaftor or Elexacaftor/Tezacaftor/Ivacaftor Treatment in Patients with CF-Associated Liver Involvement. Single measures of AST (A,G), ALT (B,H), GGT (C,I), Platelets (D,J), APRI (E,K), and GPR (F,L) before and during treatment with CFTR modulator therapy. CF-associated liver involvement was defined as having APRI ≥ 0.345 or GPR ≥ 0.403 on at least two occasions at least three months apart prior to starting CFTR modulator therapy. Individual laboratory values were analyzed and plotted as black circles. Individual patient trends are shown in black lines, and cohort averages are shown in red lines. P values are indicated for pairwise comparisons of means.

### Sub-analysis of patients with pre-existing liver involvement treated with elexacaftor/texacaftor/ivacaftor

We next sought to understand why we observed improved GGT, APRI, and GPR in those patients with underlying liver involvement treated with lumacaftor/ivacaftor but not elexacaftor/tezacaftor/ivacaftor. We examined the laboratory values of seven patients treated with lumacaftor/ivacaftor who were subsequently treated with elexacaftor/tezacaftor/ivacaftor and who had evidence of liver involvement before starting lumacaftor/ivacaftor. To first determine if it was related to the longer duration of treatment with lumacaftor/ivacaftor, we controlled for the treatment time with each drug by analyzing laboratory values obtained ∼12 months after starting each therapy (average (SD) 13.1 ± 4.2 months for lumacaftor/ivacaftor vs. 14.3 ± 4.8 months for elexacaftor/tezacaftor/ivacaftor).

While these patients showed improvement in AST, ALT, GGT, and GPRI on lumacaftor/ivacaftor, similar to the liver involvement cohort in Figure 1, they did not show any further improvements on elexacaftor/tezacaftor/ivacaftor (Figure 2). In fact, ALT worsened with elexacaftor/tezacaftor/ivacaftor. Sub-analysis of patients with underlying liver involvement who did not receive prior modulator therapy before starting elexacaftor/tezacaftor/ivacaftor (n = 6) did not identify changes in LFTs or liver fibrosis indices aside from a decrease in platelets (P = 0.030, Supplemental Figure 2). Together, these results suggest that while lumacaftor/ivacaftor improves surrogate markers of liver injury in patients with underlying evidence of involvement, elexacaftor/tezacaftor/ivacaftor does not appear to have the same effect, at least within the timeframe studied in this patient cohort.

**Figure 2.**
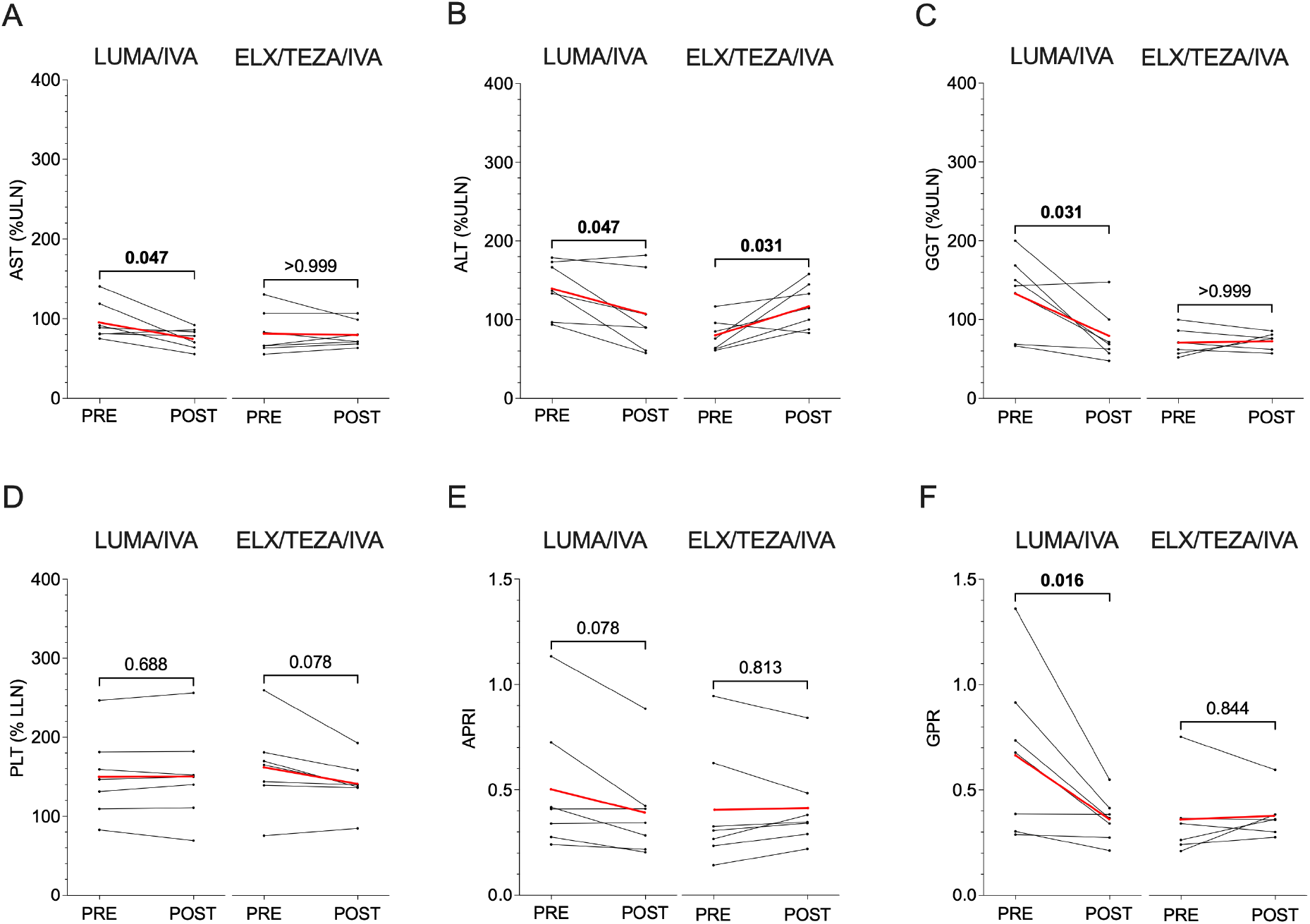
Changes in Liver Function Tests and Liver Fibrosis Indices During Lumacaftor/Ivacaftor (LUMA/IVA) followed by Elexacaftor/Tezacaftor/Ivacaftor (ELEX/TEZA/IVA) Treatment in CF Patients with Liver Involvement. AST (A), ALT (B), GGT (C), and Platelets (D) were measured, and APRI (E) and GPR (F) were calculated in the same patients before and during treatment with each CFTR modulator therapy. Individual laboratory values were analyzed and plotted as black circles. Individual patient trends are shown in black lines, and cohort averages are shown in red lines. P values are indicated for pairwise comparisons of means.

## DISCUSSION

With lung failure the primary cause for mortality in CF, clinical trials examining lumacaftor/ivacaftor and elexacaftor/tezacaftor/ivacaftor focused on changes in pulmonary function (1, 2). However, cirrhosis with portal hypertension is a significant contributor to both morbidity and mortality in CF, especially in children where its peak onset occurs. Because patients with liver disease were excluded from lumacaftor/ivacaftor and elexacaftor/tezacaftor/ivacaftor clinical trials, data on the impact of these drugs on liver health is scarce. Our observations that lumacaftor/ivacaftor improves LFTs and liver fibrosis indices in CF agree with a recent single center study by Drummond et al (11). Of note, neither of these cohorts was examined prior to starting therapy with ultrasound or MRI elastography, which is increasingly being used in conjunction with LFTs to non-invasively quantify fibrosis. In our large academic CF center, the use of ultrasound was not routinely performed for liver disease screening at the time of lumacaftor/ivacaftor approval. While it was being routinely performed prior to elexacaftor/tezacaftor/ivacaftor approval, the COVID-19 pandemic prevented many individuals from receiving a routine ultrasound prior to starting elexacaftor/tezacafttor/ivacaftor. With these caveats in mind, both cohorts likely consisted of those with mild-moderate liver involvement as assessed by biomarkers of fibrosis which correlate well with imaging findings (9). It is unclear if our findings are limited to those without advanced liver cirrhosis and/or portal hypertension, but the greatest opportunity to reverse disease likely lies prior to the development of multilobular cirrhosis.

To date, ours is the first study to report on the potential impact of elexacaftor/tezacaftor/ivacaftor on liver fibrosis indices. Given its greater efficacy to improve lung function compared to lumacaftor/ivacaftor, we hypothesized that we may see even better improvement in liver biomarkers than lumacaftor/ivacaftor. But we did not. In our liver involvement cohorts, both FEV1 and BMI percentile improved during treatment with elexacaftor/tezacaftor/ivacaftor, arguing that the lack of improvement in LFTs was not due to treatment failure or substantial medication noncompliance. Since all but one patient on lumacaftor/ivacaftor had normalization of GGT and GPR, sensitive markers of early liver fibrosis (9), this may have obscured our ability to detect any subsequent change with elexacaftor/tezacaftor/ivacaftor. The PROMISE study (Prospective Study to Evaluate Biological and Clinical Effects of Significantly Corrected CFTR Function) includes evaluation of liver function on elexacaftor/tezacaftor/ivacaftor and may provide further clarity on this question. Our data suggest it may be important to consider prior CFTR modulator exposure when analyzing the PROMISE data.

In summary, we have shown that lumacaftor/ivacaftor improves liver function studies and liver fibrosis indices in patients with pre-existing liver abnormalities. Lumacaftor/ivacaftor may represent the first drug to reverse CF-associated liver fibrosis, but larger multicenter studies with imaging and/or histology are required to validate these findings.

## Data Availability

All data produced in the present study are available upon reasonable request to the authors

## Abbreviations

CFTR: cystic fibrosis transmembrane conductance regulator
CF: cystic fibrosis
AST: aspartate aminotransferase
ALT: alanine aminotransferase
GGT: gamma-glutamyl transferase
APRI: AST-to-Platelet Ratio Index
GPR: GGT-to-Platelet Ratio
ULN: upper limit of normal
LLN: lower limit of normal
CALIPER: Canadian Laboratory Initiative on Pediatric Reference Intervals
SD: standard deviation
LFTs: liver function tests
US: ultrasound.

---

**Supplemental Figure 1.**
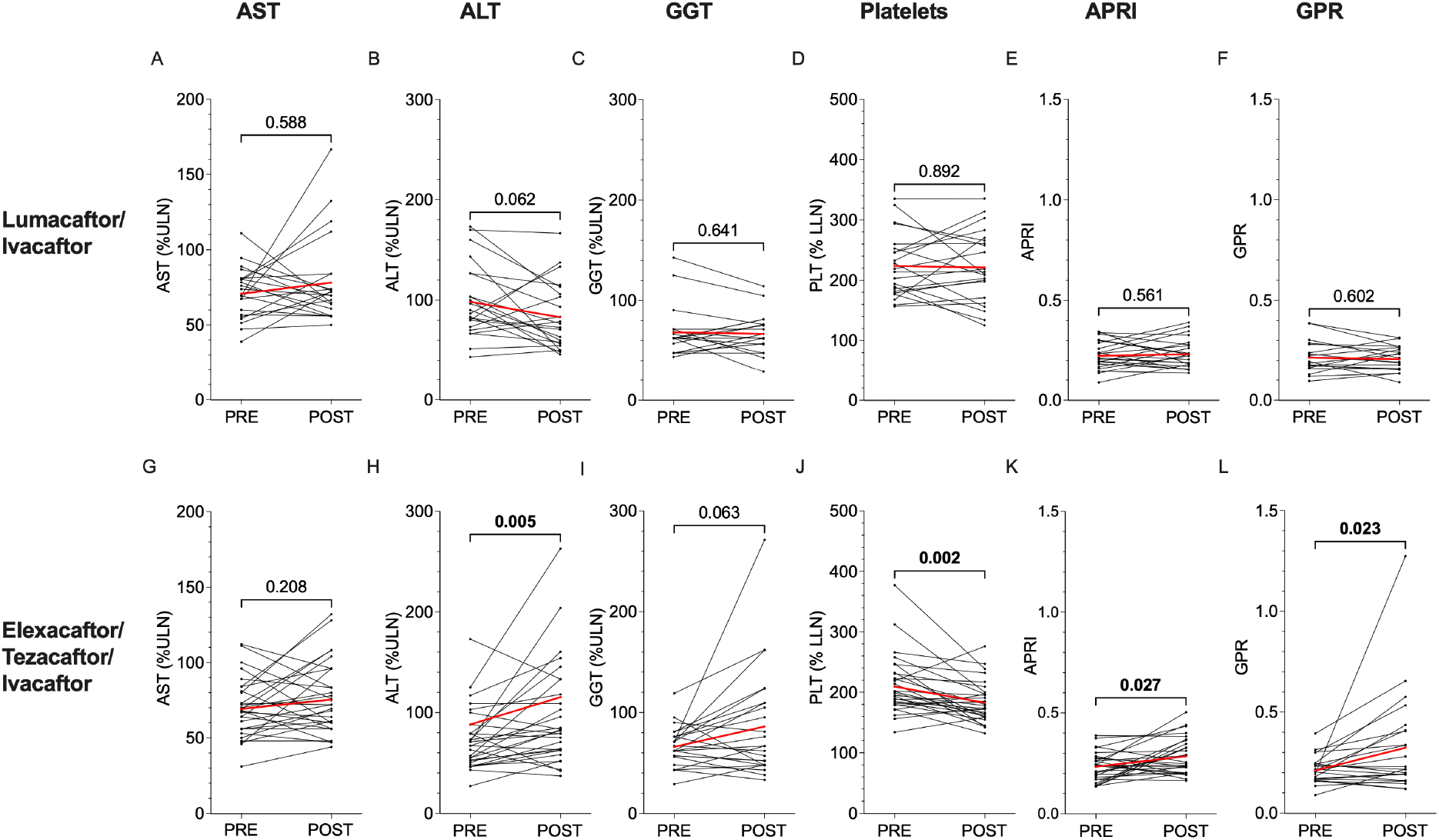
Changes in Liver Function Tests and Liver Fibrosis Indices During Lumacaftor/Ivacaftor or Elexacaftor/Tezacaftor/Ivacaftor Treatment in Patients without CF-Associated Liver Involvement. Single measures of AST (A,G), ALT (B,H), GGT (C,I), Platelets (D,J), APRI (E,K), and GPR (F,L) before and during treatment with CFTR modulator therapy. CF-associated liver involvement was defined as having APRI ≥ 0.345 or GPR ≥ 0.403 on at least two occasions at least three months apart prior to starting CFTR modulator therapy. Individual laboratory values were analyzed and plotted as black circles. Individual patient trends are shown in black lines, and cohort averages are shown in red lines. P values are indicated for pairwise comparisons of means.

**Supplemental Figure 2.**
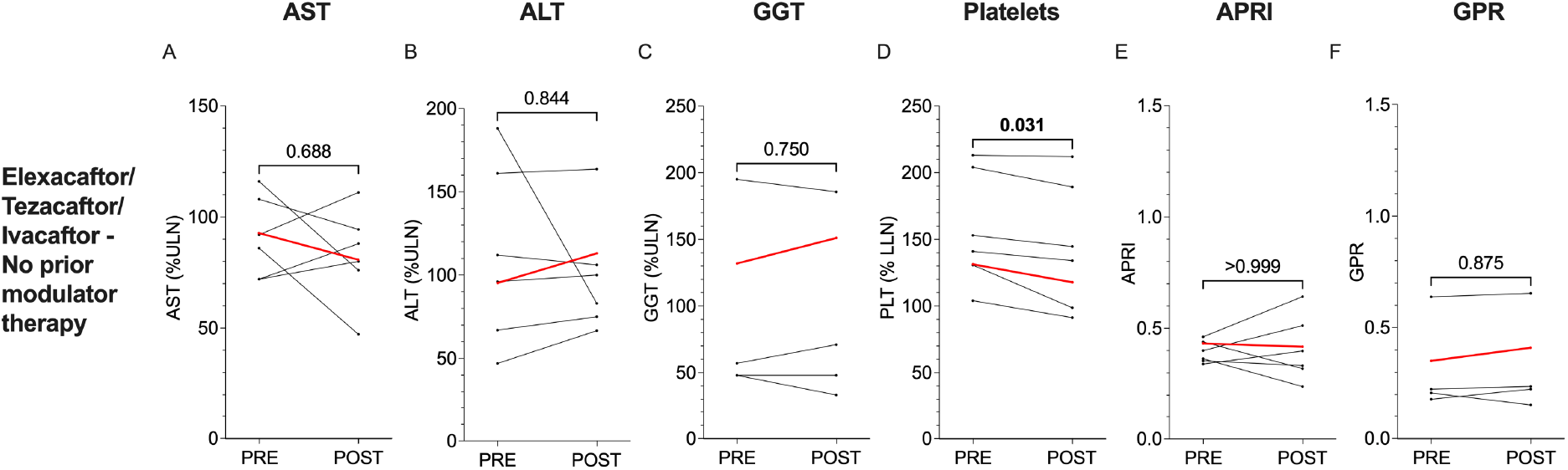
Changes in Liver Function Tests and Liver Fibrosis Indices During Elexacaftor/Tezacaftor/Ivacaftor Treatment in Patients With CF-Associated Liver Involvement Not Treated with Prior Modulator Therapy. Single measures of AST (A), ALT (B), GGT (C), Platelets (D), APRI (E), and GPR (F) before and during treatment with CFTR modulator therapy. CF-associated liver involvement was defined as having APRI ≥ 0.345 or GPR ≥ 0.403 on at least two occasions at least three months apart prior to starting CFTR modulator therapy. Individual laboratory values were analyzed and plotted as black circles. Individual patient trends are shown in black lines, and cohort averages are shown in red lines. P values are indicated for pairwise comparisons of means.

